# Health-related quality of life with herpes zoster: systematic review and analysis of individual patient data from 9 cohorts

**DOI:** 10.1101/2025.04.28.25326561

**Authors:** Chu-Chang Ku, Alicia Rosello, Peter Hobbelen, Koen B. Pouwels, Judith Breuer, Adam Gater, Kelly D. Johnson, Albert J.M. van Wijck, Melanie Drolet, Stuart Carroll, Marc Brisson, Marc Baguelin, Nick Andrews, Albert Jan van Hoek, Nicholas G. Davies, Mark Jit

## Abstract

**Background:** Herpes zoster (HZ), or shingles, is a painful rash caused by reactivation of latent varicella zoster virus infection. To better inform health economic evaluation of interventions against HZ, we synthesised evidence from studies measuring the health-related quality of life (HRQoL) of patients with HZ using the EuroQol EQ-5D instrument.

**Methods:** We systematically reviewed the scientific literature up to 31st October 2024 to identify all studies of HRQoL with HZ from PubMed and Web of Science. We obtained individual-level data from cohorts measuring HRQoL using the EQ-5D. We fit a survival model to the duration of the disutility period, and a mixture distribution to capture changes in HRQoL during the disutility period, to estimate the quality-adjusted life years (QALY) lost due to HZ by age.

**Results:** We identified 22 studies reporting a measure of HRQoL with HZ. Of these, 13 studies, covering 16 patient cohorts, measured HRQoL using the EQ-5D tool. We included individual patient data from 9 of these 16 cohorts (1,345 patients) in our quantitative synthesis. We estimated the disutility period for HZ to range between 121 (95% credible interval [CrI]: 112-134) days for 50-year-olds to 193 (163-226) days for 90-year-olds. We estimated the QALY loss due to an HZ episode to range from 0.095 (0.086 – 0.104) at 50 years to 0.146 (0.124 – 0.172) at 90 years of age.

**Conclusions:** This study is the first to derive a QALY loss estimate for HZ through a systematic review of the literature and analysis of individual patient data. Longer follow-up studies for HZ patients will be important in the future to robustly capture the effect of long-term sequelae and may potentially lead to a higher estimated QALY loss.

## Introduction

Herpes zoster (HZ), or shingles, is caused by the reactivation of latent varicella zoster virus infection. The incidence of HZ ranges from 5.2 to 10.9 cases per 1,000 person-years in individuals ≥50 years of age in high-income countries ^1^. In most cases, HZ involves a painful and/or pruritic blistering rash. The duration of the rash is about 7-10 days while the pain can persist for months ^2^. Postherpetic neuralgia (PHN) is the most common complication of HZ cases (occurring in 5–30% of HZ cases) ^2^, usually defined by persistent pain lasting 3 months or longer. Beyond the pain, patients with PHN may suffer from depression and social disconnection ^3^. Other complications from HZ include zoster ophthalmicus (5% of HZ cases), bacterial superinfection of the lesions, facial palsy, paresis, pneumonia, and encephalitis ^2^.

Cost-utility analysis in health technology assessment (HTA) is essential in health-related decision-making. As a part of cost-utility analysis, the reductions in health-related quality of life (HRQoL) resulting from health issues are used to evaluate the associated health burden. Several observational studies and clinical trials have measured HRQoL losses associated with cases of HZ, using a variety of HRQoL surveys and study designs. However, to date there has been no synthesis of the results of these various studies, and as such we lack a comprehensive understanding of the quantifiable health losses associated with HZ cases, and of how this burden evolves over the course of the illness.

Here, we systematically reviewed the QALY loss due to HZ from a disease dynamic perspective. We quantitatively synthesised measurements of HRQoL measured by EuroQol’s EQ-5D instrument to characterise the distribution of HRQoL over time since HZ-associated rash onset from a disease-dynamic viewpoint. We also derived estimates of the total QALY loss per HZ case as a function of patient age and under different baseline health assumptions to support further HZ-related HTAs.

## Methods

### Search strategy

Studies describing the HRQoL for people with HZ were identified by searching PubMed and Web of Science. We searched for (“shingles” OR “zoster”) AND (“burden” OR “QALY” OR “utility” OR “quality of life”) in abstracts or titles among English-language, peer-reviewed studies published from 1^st^ January 2008 to 31^st^ October 2024. We aimed to identify all publications reporting novel HRQoL estimates for HZ patients among the general population in observational studies. We excluded studies restricted to specific types of HZ (e.g. HZ with PHN only), sub-populations (e.g. immunocompromised patients), or healthcare settings (e.g. hospital-treated cases). AR and PH conducted an initial assessment, reviewing titles, abstracts and full texts for those studies published before 2017 from PubMed. Then ND and CK finalised the review protocol and conducted a second round accordingly. The same authors reviewed full texts to collect studies that met the inclusion and exclusion criteria. The identified studies which reported using the EQ-5D tool to measure HRQoL were selected for quantitative synthesis to estimate QALY loss due to HZ. Study authors were contacted to obtain patient-level EQ-5D data by time since HZ rash onset and patient age at rash onset.

### Inclusion and exclusion criteria

Studies that provided novel estimates of HRQoL for patients with HZ were included in the review. We focus on studies that collected original HRQoL data for patients that would be representative of the general population presenting to primary care with HZ. We also included the following types of studies for a snowballing screening: (1) studies of cost-effectiveness/cost-utility analysis, and burden estimates and model projections for inputs for HRQoL with HZ if applicable, and (2) review studies that may have included previously-published primary research articles with HRQoL measurements for HZ. Case studies and studies based on specialised or tertiary healthcare were excluded; trial-based studies in which patients were recruited via active surveillance were not included. From full-text assessment, we excluded studies that did not have new HRQoL estimates or that did not collect temporal information relative to the onset dates. Studies that only measured HRQoL in a specific type of patient with HZ (e.g. HZ with PHN) and studies that did not report quantitative scores were excluded. When multiple studies were based on the same data sets, we included the studies which we judged to include the most detail relevant for our study. In one case (the MASTER study across eight countries) we kept both the earlier summary paper describing the results across all countries, and later papers which focused on three of the nine countries in the study in greater detail.

### Data extraction of EQ-5D scores

The EQ-5D survey asks participants to record whether they have no problems, some problems, or severe problems across five dimensions of HRQoL. These are then transformed using country-specific valuation sets, which yield a single number representing overall HRQoL, referred to as the EQ-5D score. An EQ-5D score of 1 indicates perfect health; an EQ-5D score of 0 indicates no quality of life or death; and negative EQ-5D scores representing states “worse than death” are also possible ^4^. From among the larger group of studies included in our review, we identified a subset of studies reporting EQ-5D scores for patients with HZ. We contacted the authors of these studies to obtain individual patient-level data comprising: EQ-5D scores measuring HRQoL for each patient, the number of days since rash onset at which each HRQoL measurement was taken (or the scheduled survey time, for those studies that did not record the survey time), patient age, and baseline EQ-5D scores prior to HZ onset, when available. We only kept subjects with at least four EQ-5D measurements after HZ rash onset. For studies which did record the time-point (date or days after the first visit) of each measurement, we excluded data points for which the time-point was missing.

### Statistical analysis

The individual patient-level data we synthesize comprise a series of HRQoL measurements over time for each patient. We fitted statistical models to characterise two aspects of HRQoL of HZ patients from these data: (1) the duration of the disutility period associated with HZ and (2) the distribution of EQ-5D scores during the disutility period. We conducted exploratory analyses, which are detailed in the Supporting Material 2, to select our statistical models.

The disutility period is defined as the duration (in days) from rash onset until the HZ patient no longer has a measurable loss in HRQoL associated with HZ. Patients in the included studies sometimes reported perfect health at earlier timepoints, but less than perfect health at later timepoints. For this reason, we assumed that the end of a given patient’s disutility period occurred between the patient’s last observation of imperfect health (EQ-5D score < 1) and the next observation of perfect health (EQ-5D score = 1). If a given patient’s last data point was not at perfect health, we considered the end of the disutility period to be censored. A Cox proportional hazards function was employed to parametrise the age dependence of the time to recovery. The baseline survival function were selected among Weibull, gamma, exponential, log-norm, and Gompertz distributions. We selected the best-fitting baseline model and function for age dependence based on Watanabe-Akaike Information Criteria and leave-one-out cross-validation. (details in Supporting Material 2) In additional to the perfect health as endpoints, we also replicated the analyses with alternative baselines of pre-HZ EQ-5D, retrospectively assessed in some MASTER studies^5–8^ and country specific population norms.

Within the disutility period, we modelled patient-reported EQ-5D scores considering patient age and time since rash onset as potential covariates. Our exploratory analysis suggested that reported EQ-5D scores appeared to fall into three different clusters. Therefore, we proposed a mixture regression model with three latent classes representing patients who reported being (1) temporarily well (EQ-5D score = 1), (2) in mild discomfort, and (3) in severe discomfort.

Specifically, we assumed that the probability of an individual experiencing HZ symptoms belonging to latent class *i* ∈ {1, 2, 3} at time *t* for age *a* is a function *f*(*i*,*t*,*a*) with ∑*_i_ f*(*i*,*t*,*a*) = 1; for each cluster we explored models with different forms of *f*(*i*,*t*,*a*) for age- and time-dependence, as well as random effects by study cohort to select the best-fitting model (details in Supporting Material 2).

### Calculation of QALY loss due to HZ

The QALY loss for HZ was defined as the cumulative loss of HRQoL from a baseline during the HZ disutility period. We aligned each baseline with the endpoints that defined the disutility period. We applied time preferences of 0%, 1.5% and 3.5% in the calculation to accommodate different decision-making settings. The details of the formulation can be found in Supporting Material 2. We characterised uncertainties of 95% credible intervals by summarizing 1,000 draws from the posterior distributions yielded by our statistical model fits.

All analyses were performed using R version 4.4.3 ^9^ and the source code is available online at [https://github.com/TimeWz667/HPRU_HZQoL].

## Results

After deduplication, we screened the titles and abstracts of 1,208 potentially relevant studies, including 101 in the full-text assessment (Fig. 1). Twenty-five studies met the selection criteria; there were 19 studies from the initial search before snowballing; then 6 were identified from the reviews or modelling studies. After removing 3 studies which potentially duplicate others (i.e., studies based on the same surveys), we included 22 studies in the qualitative synthesis (Table S1).

**Fig 1.**
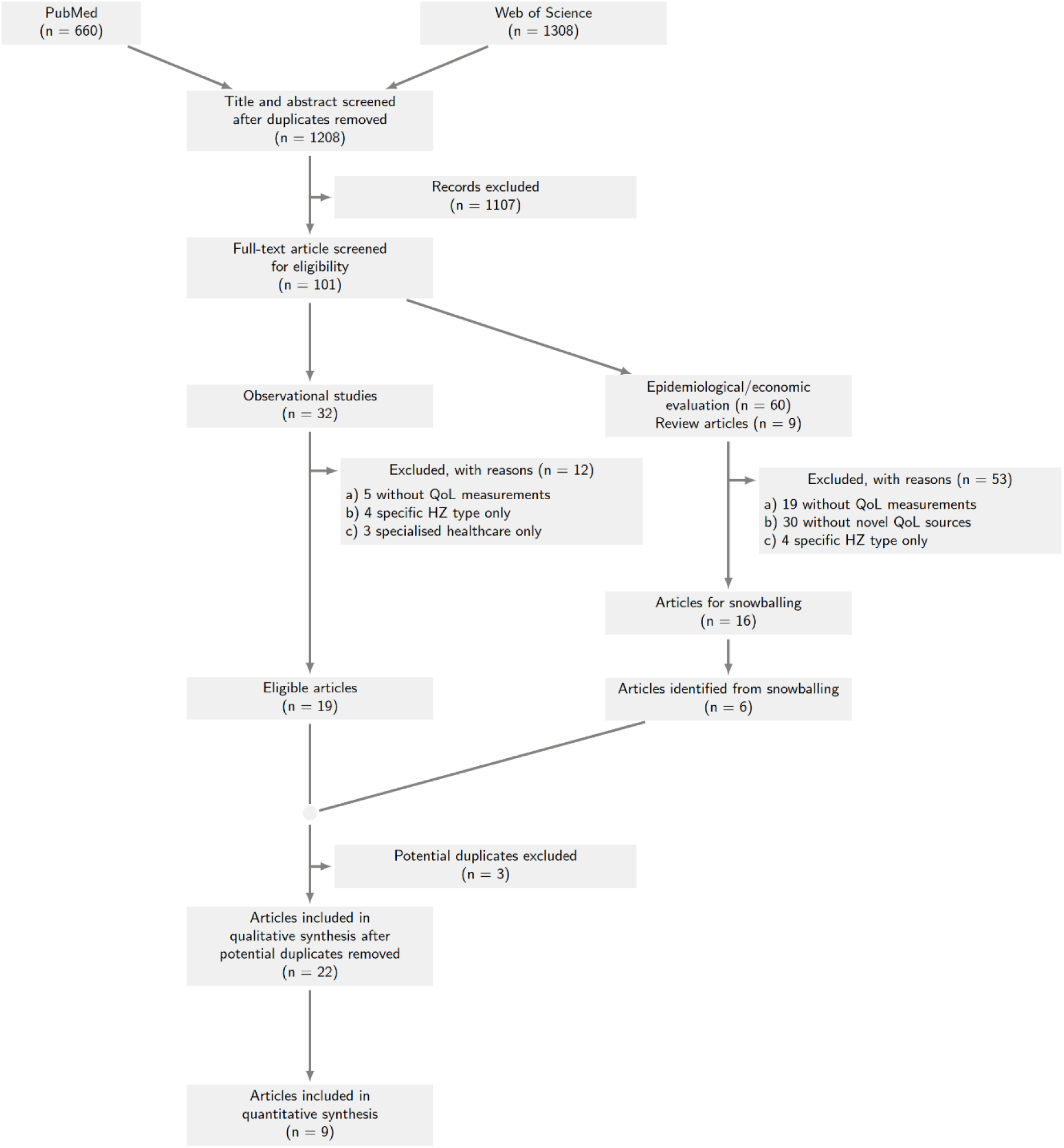
PRISMA flow diagram.

### Study characteristics

The included studies comprised single-country studies based in the United Kingdom ^10,11^, the United States ^12–14^, France ^15–17^, Italy ^18–21^, Germany ^22^, Greece ^23^, the Netherlands ^24^, Spain ^25^, Canada ^26^, Japan ^27^, South Korea ^28^, Taiwan ^29^, and Colombia ^30^, and one multi-country study based in Argentina, Brazil, Costa Rica, Mexico, Canada, Korea, Taiwan and Thailand ^31^. Chronologically, there were five studies published in 2008 or earlier (identified through snowballing ^10,12–14,18^), 15 published between 2009 and 2019 ^11,15–17,19–24,26–29,31^, and two articles published in 2020 or later ^25,30^.

Across the 22 included studies, HZ-associated disutility was measured using a variety of different instruments, some assessing quality of life along both physical and mental health dimensions and others focusing on physical pain or mental ill health exclusively (Table S1). Among the general quality of life instruments, 13 articles reported EQ-5D measurements ^10,11,14,21,22,24–31^, nine studies used the SF-12 ^12,15–17,19,20,24,26,27^ two used the EQ visual analogue scale (EQ-VAS) ^12,22,31^, one used the Zoster Impact Questionnaire ^12^, and one used time trade-off ^13^. Among the pain-specific instruments, six studies used the Zoster Brief Pain Inventory ^12,21,25,27,30,31^ and two used the McGill Pain Questionnaire Present Pain Intensity instrument ^12,27^. Focusing on mental health, three studies used the Hospital Anxiety and Depression Scale ^15–17^ and one study^23^ reported Short Anxiety Screening Test scores ^23^.

### Quantitative synthesis

From among the 13 included studies reporting EQ-5D scores and therefore eligible for the quantitative synthesis, 16 patient cohorts were described. Two of these studies ^11^ ^32^ had no patients with 4 or more HRQoL measurements taken, and so did not contribute any data to the quantitative synthesis. Data were not available for five studies ^21,22,25,27,33^. From the six remaining studies, we obtained individual patient data for 9 patient cohorts from the study authors (Table 1). Scott et al.^10^, and Van Wijck et al.^24^ provided individual patient data from their studies. Rampakakis et al. ^31^ pooled several patient cohorts, restricted to patients engaged in full or part-time work, using similar protocols (referred to as the MASTER studies); three of these cohorts were described separately by Song et al.^28^, Tsai et al.^29^ and Drolet et al.^26^. In the analysis, we removed duplicated samples to prevent double counting. Individual patient data was obtained from 7 of 8 MASTER patient cohorts; the authors cannot share data from their cohort of patients recruited in Brazil.

**Table 1.** List of articles included in the quantitative synthesis.

|  | **MASTER studies from Rampakakis et al. <sup>31</sup>** | **MASTER study from Drolet et al. <sup>26</sup>** | **MASTER study from Song et al. <sup>28</sup>** | **MASTER study from Tsai et al. <sup>29</sup>** | **Scott et al. <sup>10</sup>** | **van Wijck et al. <sup>24</sup>** |
| --- | --- | --- | --- | --- | --- | --- |
| **Country (N. patients included in synthesis)** | Argentina (53), Costa Rica (7), Mexico (39), Thailand (162) | Canada (263) | Korea (89) | Taiwan (130) | UK (55) | Netherlands (547) |
| **Study period** | 2005-2012 | 2005-2006 | 2009-2010 | 2008-2009 | 2002-2003 | 2010-2014 |
| **Age** | ≥50 | ≥50 | ≥50 | ≥50 | Any | ≥50 |
| **Enrolment diagnosis** | HZ rash or HZ-associated pain | Physician-confirmed diagnosis of HZ (103/258 PCR confirmed) | Physician-confirmed diagnosis of HZ | HZ rash or HZ-associated pain | GP referral and confirmation by PCR | Acute eruptive zoster ≤7d after rash onset |
| **Exclusion criteria** | Any medical condition that could interfere with evaluations required by the study, and patient and/or family member or primary caregiver refusal to sign informed consent. | Previously vaccinated for HZ, unable to complete questionnaires in French or English | Unable to complete questionnaires in Korean | Presence of any medical condition that could interfere with evaluations |  | Language |
| **Timing measurement s HRQoL** | Initial assessment and 7d, 14d, 21d, 1m, 2m, 3m, 4m, 5m and 6m after rash onset | Initial assessment and 7d, 14d, 21d, 1m, 2m, 3m, 4m, 5m and 6m after initial assessment | Initial assessment and 7d, 14d, 21d, 1m, 2m, 3m, 4m, 5m and 6m after initial assessment | Initial assessment and 7d, 14d, 21d, 1m, 2m, 3m, 4m, 5m and 6m after initial assessment | Initial assessment and 1m, 3m and 6m following enrolment | Initial assessment, 14d, 1m, 3m, 6m, 9m, and 1y |

1,345 patients, with 9,810 observations in total, were included in the statistical analysis. The number of measures ranged from 4 to 10 per patient, and 452 patients (34%) had at least one observation made more than 365 days after rash onset. The median follow-up time was 191 days, and the median time to the last observation of less than perfect health was 38 days. 309 patients (23%) had the full number of observations according to the study design. At their last observations, 292 patients (22%) reported less than perfect health; this is the proportion of patients for whom the disutility period is considered censored in our analysis. The mean EQ-5D score reported across all patients was 0.59 for the first two weeks after rash onset if they were not in perfect health.

For the duration of the disutility period (Fig. 2A), we fitted a Cox proportional hazards model with parametric baseline hazard function. An exponential distribution was selected for the baseline hazard function from Weibull, gamma, exponential, log-normal, and Gompertz distributions. After variable selection, the final model included a term for patient age but no term for study cohort. We found older patients took longer to recover from HZ. The median projected disutility period (Fig. 2B, C) for people aged 50 is 121 (95% CrI: 112-134) days; for people aged 70, 153 (95% CrI: 163-226) days; and for people aged 90, 193 days.

**Fig 2.**
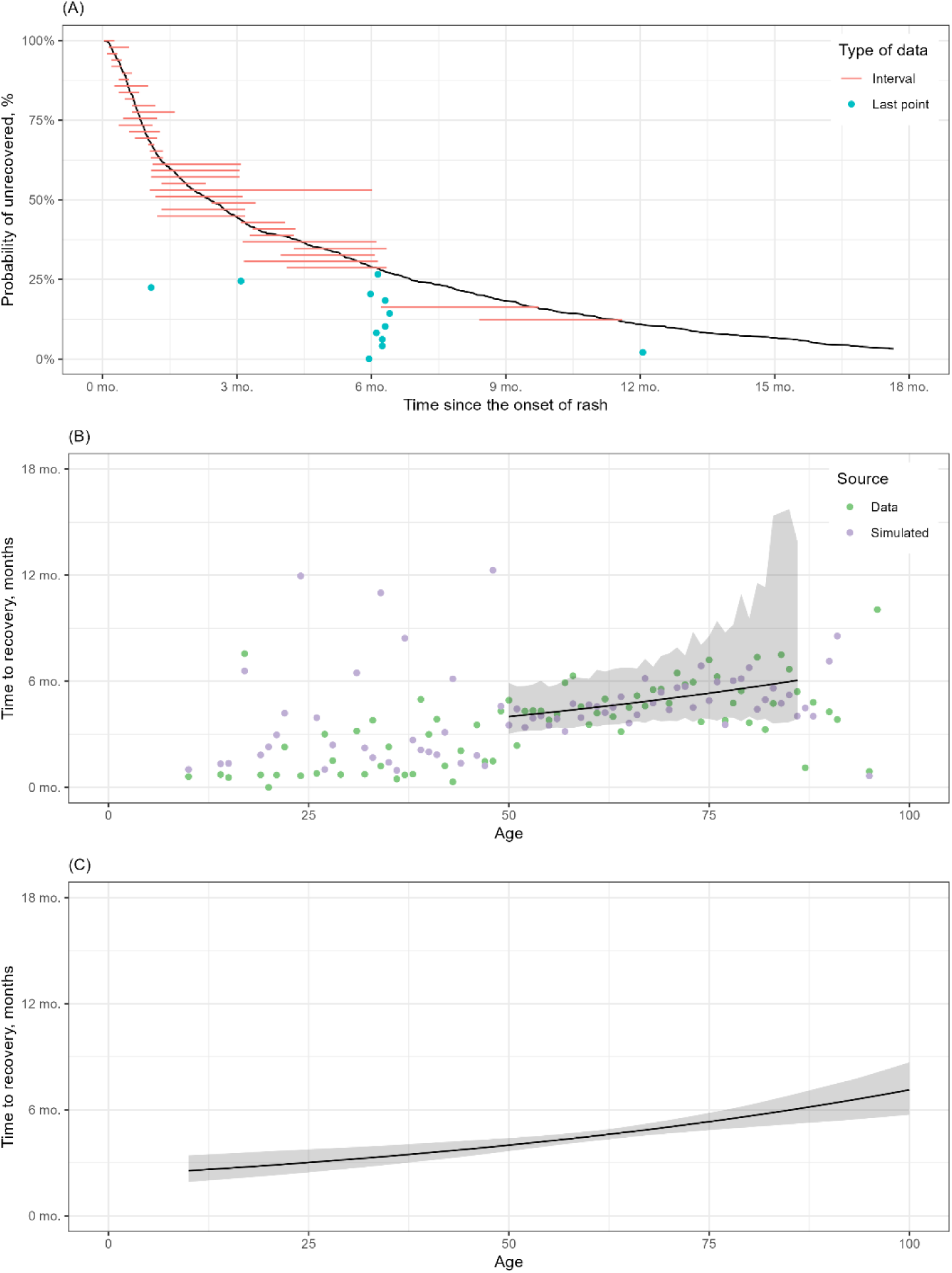
Disutility period. (A) Time to the end of disutility period (survival function) with data samples. Red ranges are for data with intervals framing recovery time, and blue points are data with last observations if recovery not observed in the data; 100 data points presented for clarity. (B) Average disutility period by age. Red points for mean of imputed recovery time, blue points for mean of simulated recovery time given the same population size as data. (C) 95% credible intervals of disutility periods by age.

Regarding HRQoL loss during the disutility period, there are three clusters in which EQ-5D scores fall: severe discomfort, mild discomfort, and temporarily well (Fig. 3A). Across all studies, 444 (35%) patients reported at least one EQ-5D score in the severe discomfort cluster; 63 (4.9%) patients reported did so 90 days after rash onset or later; 341 (27%) patients were “temporarily well” at least once. The mean EQ-5D score in the severe discomfort cluster 0.10 (95% CrI: −0.481 – 0.378) and in the mild discomfort cluster is 0.74 (95% CrI: 0.52 – 0.85). Disregarding the cohort-specific random effects, the predictions of the EQ-5D score are 0.064 (95% CrI: 0.049 – 0.078) and 0.722 (95% CrI: 0.718 – 0.726) respectively. The proportion of severe scores decreased from 16% in the first three months after rash onset to lower than 10% thereafter (Fig. 3B). No significant age-specific patterns within the two clusters were identified: the proportion of observations in each cluster varied by time until 30 days after rash onset.

**Fig 3.**
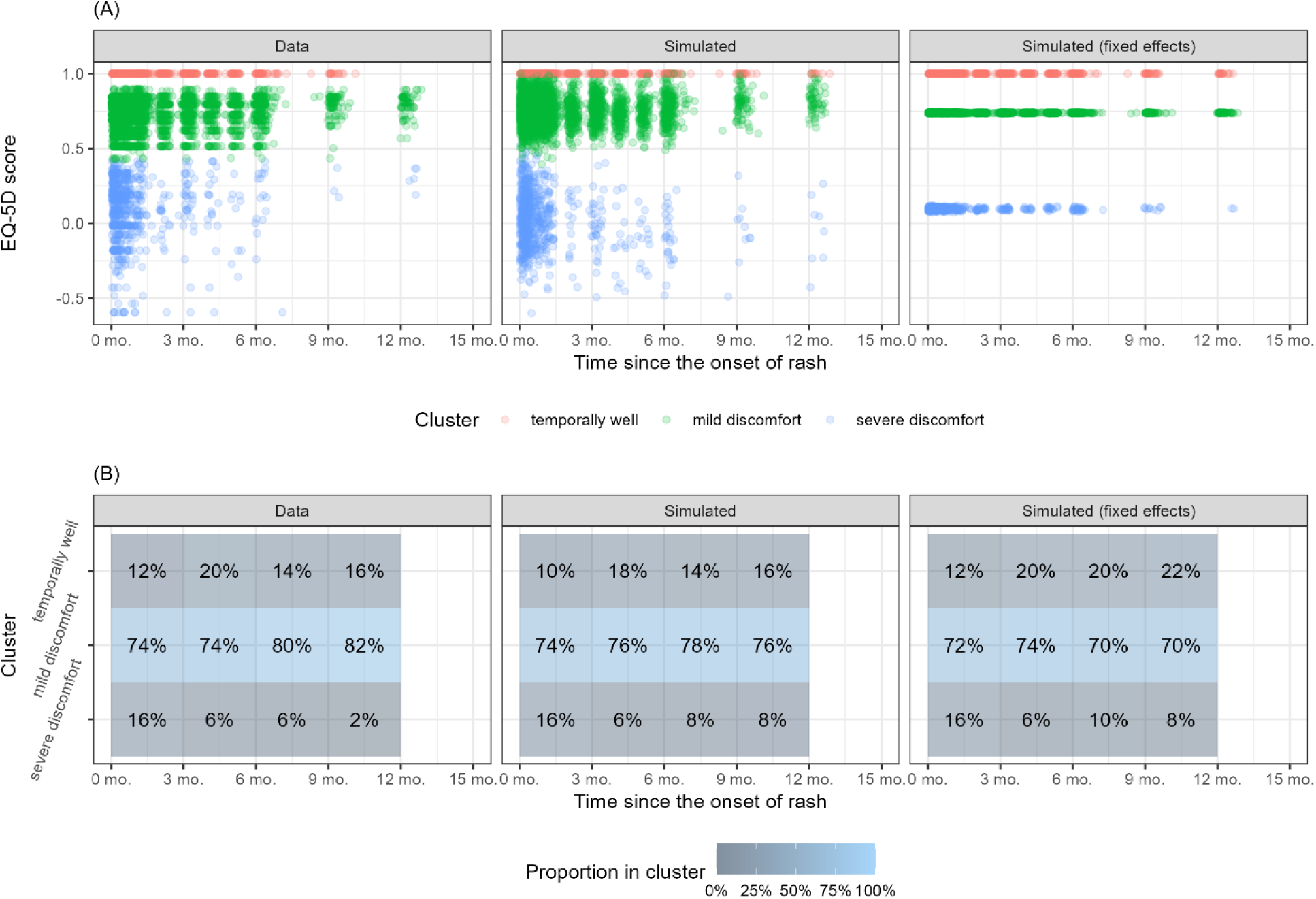
EQ-5D scores. Left panel for the original data, middle panel for simulated data, and right panel for simulated data without the random effects related study cohorts removed. Re points indicate the temporarily well, green points and blue points indicate the mild and severe discomfort respectively. The simulated data points follow the same age- and time- distribution.

### QALY loss due to HZ

Taking perfect health as the baseline, the total QALY loss resulting from a case of HZ is 0.095 QALYs (95% CrI: 0.086 – 0.106) at 50 years, increasing to 0.146 QALYs (95% CrI: 0.124 – 0.172) at 90 years (Table 2). With the per-HZ EQ-5D baselines, only about a half of QALY loss were estimated across all ages. However, applying the population norms according the study settings, the QALY loss decreases with increasing age, with a loss of 0.046 QALYs (95% CrI: 0.042 – 0.051) per HZ case at 50 years, decreasing to 0.037 QALYs (95% CrI: 0.031 – 0.044) at 90 years (Fig. 4B). These results correspond to an assumed no discounting for time preference; varying this rate to 1.5% or 3.5% annually changes these results by less than 0.002 QALY across all scenarios.

**Table 2.** Duration of disutility period and QALY loss due to HZ by different assumptions of HZ-free baseline.

| HZ-free baseline | Age | Disutility period, days (95% CrI) | HZ-related HRQoL loss, QALY (95% CrI), no discounting | HZ-related HRQoL loss, QALY (95% CrI), 1.5% discounting | HZ-related HRQoL loss, QALY (95% CrI), 3.5% discounting |
| --- | --- | --- | --- | --- | --- |
| Perfect health | 50 | 121 (112 - 134) | 0.095 (0.086 - 0.106) | 0.094 (0.086 - 0.105) | 0.094 (0.085 - 0.105) |
|  | 60 | 136 (128 - 146) | 0.105 (0.098 - 0.115) | 0.105 (0.097 - 0.114) | 0.104 (0.096 - 0.113) |
|  | 70 | 153 (142 - 166) | 0.118 (0.108 - 0.129) | 0.117 (0.107 - 0.128) | 0.116 (0.106 - 0.127) |
|  | 80 | 172 (153 - 192) | 0.131 (0.116 - 0.148) | 0.130 (0.116 - 0.146) | 0.129 (0.115 - 0.145) |
|  | 90 | 193 (163 - 226) | 0.146 (0.124 - 0.172) | 0.145 (0.123 - 0.170) | 0.143 (0.122 - 0.168) |
| Population norm<sup>†</sup> | 50 | 116 (106 - 126) | 0.050 (0.045 - 0.055) | 0.050 (0.045 - 0.055) | 0.050 (0.045 - 0.055) |
|  | 60 | 122 (114 - 130) | 0.046 (0.042 - 0.051) | 0.046 (0.042 - 0.050) | 0.046 (0.042 - 0.050) |
|  | 70 | 128 (119 - 138) | 0.042 (0.038 - 0.047) | 0.042 (0.038 - 0.047) | 0.042 (0.038 - 0.046) |
|  | 80 | 135 (121 - 150) | 0.039 (0.034 - 0.044) | 0.039 (0.034 - 0.044) | 0.038 (0.034 - 0.044) |
|  | 90 | 143 (121 - 166) | 0.037 (0.031 - 0.044) | 0.037 (0.031 - 0.043) | 0.037 (0.031 - 0.043) |
| Pre-HZ baseline | 50 | 90 (81 - 100) | 0.057 (0.044 - 0.069) | 0.057 (0.044 - 0.069) | 0.057 (0.043 - 0.068) |
|  | 60 | 104 (96 - 114) | 0.064 (0.050 - 0.078) | 0.064 (0.050 - 0.078) | 0.064 (0.050 - 0.077) |
|  | 70 | 121 (109 - 134) | 0.071 (0.053 - 0.088) | 0.070 (0.053 - 0.088) | 0.070 (0.052 - 0.087) |
|  | 80 | 141 (121 - 164) | 0.077 (0.055 - 0.100) | 0.076 (0.055 - 0.100) | 0.076 (0.055 - 0.099) |
|  | 90 | 163 (134 - 203) | 0.079 (0.053 - 0.112) | 0.078 (0.053 - 0.112) | 0.078 (0.053 - 0.111) |
| †Population norms are averages by sample size used from each study; see Supplementary Material 2 for study-specific results |  |  |  |  |  |

## Discussion

We estimated the health-related quality of life loss due to herpes zoster through a systematic review of the literature and analysis of individual patient data. Our quantitative synthesis combines data from 1,345 patients across 9 countries. We found that while older patients generally suffer from HZ for longer, their reported HRQoL during the illness episode was not significantly worse than that of younger patients. The longer disutility period for older HZ patients leads to greater QALY loss when we assume that reported EQ-5D scores capture HZ-attributable HRQoL only, i.e. using a baseline of perfect health. However, when we assume that reported EQ-5D scores capture both HZ-attributable and underlying HRQoL — i.e. using a baseline of the population norm — then since the older population has a worse baseline health status, younger HZ patients experience a greater loss in quality of life than older patients despite their shorter disutility period.

The use of perfect health as a baseline is commonly found in QALY loss assessments for HZ, such as those by Matthews et al. ^21^ and Diez-Domingo et al. ^25^. Alternatively, some national guidelines for HTA, such as those for the UK ^34^, prioritise the population norm as the baseline in order to better reflect the disease burden. Our estimation showed an opposing age-specific pattern for HZ patients aged 50 and older across these different baselines. This study is the first to quantify this feature systematically, providing three alternatives for evaluating HRQoL in HZ-related HTAs.

Earlier work by van Hoek et al.^35^ summarised pain-centred HRQoL prior to the introduction of any vaccines. Sollie et al. ^36^ conducted a systematic review of patient-reported HRQoL and synthesized results from five studies using the SF-12 instrument, but only considered the average HRQoL loss, without accounting for the duration of the disutility period or the dynamics of the HRQoL loss over time. Our study explores the HRQoL loss due to HZ from real-world settings in depth. As a result, all nine data sets included in our quantitative synthesis were from observational studies. Of note, most of the studies were conducted in high-income settings. We did not identify substantial heterogeneity among the study samples other than that associated with differences in patient age across studies, although two of the three upper-middle-income countries, Costa Rica and Thailand, showed slightly lower and slightly higher EQ-5D scores, respectively.

This analysis also aims to inform decision-making related to HZ from the healthcare system perspective. In existing economic analyses for HZ, i.e., evaluations of recombinant zoster vaccine reviewed in Giannelos et al.^37^, the cost and the utility were usually based on the incidence of HZ estimated from medical records. While trial-based QALY may capture a wider population affected by HZ, QALY loss from real-world data is more relevant for weighing medical costs and health burden on the same basis, unless underdiagnosis and care-seeking behaviours are explicitly addressed to stratify individuals who sought care for HZ and those who did not.

This study provides parameter estimates for HTA from the healthcare system perspective. Most HTAs did not model individuals who do not seek care, implicitly assuming that people with HZ unless death or early recovery occurs first. We modelled the dynamics of HRQoL by separating time to recovery and using HRQoL as the weight for the disutility period. This approach is compatible with Markov-style models and ODE systems, which are commonly employed in HTAs.The study presented a quantitative exploration of the dynamics of HRQoL after rash onset, using patient-level data rather than a meta-analysis of the original disutility estimates. We challenge two common practices in HRQoL analyses for HZ: (1) assuming a 90-day disutility period and (2) using summary metrics that overlook the heterogeneity of HRQoL losses. Only one study, Curran et al.^22^, addressed the severe pain due to HZ earlier than the formal definition of PHN by relating EQ-5D scores to Zoster Brief Pain Inventory measures. Neglecting heterogeneity might lead to an underestimation of QALY loss when assessed against a baseline, as illustrated in the final section of Supplementary Material 2. The key rationale is that most HZ patients experience pain which is not significantly worse than their baseline health status. Severely affected patients are too few in number for their experience to shift the average substantially.

Our disutility period analysis quantified that the expected duration of HZ among those aged 80 and older is longer than five months, and that the 90-day cutoff captures only about half of the data. We found a large share of patients have HZ lasting longer than 90 days who might not be classified as having PHN in other analyses. However, many HRQoL studies used a 90-day cut-off point when calculating QALY losses, e.g., Mizukami et al.^27^ and Matthew et al.^21^. That means disutility beyond 90 days is not counted, even though it contributed around 60% of the total QALY loss among patients aged 70–80 years in our simulations. Tsai et al.^29^ also revealed a long tail of moderate painfulness among people aged 50 and older, highlighting the potential value of preventive interventions for HZ. Furthermore, HRQoL in some patients was not fully captured due to the length of the follow-up in original analyses, leaving 23% of patients unrecovered by the end of the observation period. Our analysis accounted for these patients within the likelihood function when parameterising time to recovery, thereby controlling for underestimation of QALY loss from censored follow-up periods. Although it is generally agreed that most HZ cases clinically resolve within 90 days, our results suggest that future HRQoL studies should extend the observation period to improve the accuracy of QALY loss estimation.

Our study highlighted the clustering of EQ-5D scores and the dynamics of transitions between these clusters, which were not reported in the original articles. Only a small proportion of patients remained in the same cluster throughout the course of illness. We did not examine the correlation between severe disutility and a PHN diagnosis. However, the proportion of patients who ever experienced severe HRQoL loss was much higher than the reported range of PHN incidence (33% compared with 5–30% in the literature ^1,2^). Furthermore, we did not find specific difference of HRQoL before and after 90 days, the threshold commonly used to define PHN. This study cannot identify whether this is because of PHN underdiagnosis or because many cases of severe HZ were clinically insufficient to meet the PHN definition. Either explanation may imply that the incidence of PHN or the disutility associated with HZ is currently underestimated. That raises a caveat for HTAs evaluating HZ preventives. Further investigation is also needed to address the public health impact of HZ.

Rampakakis et. al. ^31^ and related studies ^26,28,29^ provided EQ-5D scores that were retrospectively collected for some patients. From those with baseline EQ-5D data, 38% of patients reported non-perfect baseline scores. Of these, 80% reported perfect EQ-5D scores at least once during follow-up. This can be regarded as a response shift, i.e. a change to how patients evaluate their own health^38,39^. From a psychological perspective, adaptation during illness and reprioritisation on quality of life domains might cause the response shift. For example, a patient suffering from impaired mobility might prioritise mobility more highly. However, within each cluster of EQ-5D scores, no consistent trends were identified across cohorts. Alternatively, from a measurement perspective, changes in responses can result from recall bias, a common limitation of retrospective surveys. Further studies with appropriate designs are needed to investigate (e.g. the then-test design). To our knowledge, there is currently no conclusive method to quantify HRQoL loss while accounting for response shift. In this study, we sought to estimate the upper bound of QALY loss by assuming a baseline of perfect health in our statistical modelling.

Supplementary Material 2 provides alternative results based on patient data with recorded baseline scores. We found that disutility period estimates were reduced by 26% in 50-year-olds and by 14% in 90-year-olds; the difference in HRQoL estimates were less than 3% across all ages. Overall, the response shift could introduce bias in QALY loss estimates, ranging from 28% in 50-year-olds to 16% in 90-year-olds.Immunocompromised individuals are at a higher risk of developing HZ and its complications ^40–42^. This population may present different QALY losses from immunocompetent individuals. The studies from which individual patient data were obtained did not exclude immunocompromised patients. Rampakakis et al.^31^ reported that 29 (6.8%) of the patients included in their analyses were immunocompromised. Scott et al. ^10^ collected this information but did not report it in their publication. Given the small number of immunocompromised patients in these studies, a sub-analysis of our data by immune status would be unlikely to yield meaningful results.

## Data Availability

The datasets generated and/or analysed during the current study are not publicly available due to limitations provided by the funders of the original studies (Sanofi Pasteur MSD, MSD) but are available from the corresponding author on reasonable request.

## Acknowledgments

We would like to thank Nikolaos Demiris and Antonio Gasparrini for their statistical advice on modelling EQ-5D data, and Gayatri Amarthalingam for helpful discussions.

## Ethics approval and consent to participate

All patients provided informed consent prior to their inclusion in each of the MASTER studies (including additional data provided in Drolet et al.^7^), the Scott et al.^43^, the Van Wijck et al.^44^, and the Gater et al.^45^. All studies were conducted as per Good Clinical Practices and the tenets of the Declaration of Helsinki. All data were collected in an anonymous fashion and as per local data protection laws, and all studies were approved by local and central Ethics Boards, as required, for each participating site. This post-hoc analysis of available data was covered in the original ethics submission.

## Authors’ contributions

CK, AR, MBa and MJ designed the study. CK, AR, ND, and PH carried out the systematic review of the literature. CK, AR, KBP, NA, AJvH and MJ contributed to the analysis. JB, AG, KJ, AvW, MD, SC, MBr provided the patient-level data on which the individual patient data meta-analysis was based. CK, AR, and ND drafted the manuscript, and all authors revised it critically for intellectual content. All authors approved the final version of the manuscript and agreed to be accountable for all aspects of the work.

## List of abbreviations

CrI: credible interval
HRQoL: health-related quality of life
HTA: health technology assessment
HZ: herpes zoster
PHN: post-herpetic neuralgia
QALY: quality-adjusted life years

---

Supplementary Material 1: PRISMA checklist

Supplementary Material 2: supporting methods and results

**Supplementary Table S1.** Characteristics of included studies.

| Author | Year | Locale | Population | Quality of life measurement | Eligible for / included<br>in the quantitative<br>synthesis | Identified by<br>snowballing |
| --- | --- | --- | --- | --- | --- | --- |
| Bouhassira et al. <sup>15</sup> | 2012 | France | ≥50 | SF-12, HADS | N |  |
| Bricout et al. <sup>19</sup> | 2014 | Italy | ≥50 | SF-12 | N |  |
| Coplan et al. <sup>12</sup> | 2004 | US | ≥60 | EQ-VAS, SF-12, McGill, ZIQ, ZBPI | N | Y |
| Curran et al. <sup>22</sup> | 2018 | Germany | ≥50 | **EQ-5D**, EQ-VAS | Y/N (not available) |  |
| Diez-Domingo et al. <sup>25</sup> | 2021 | Spain | ≥50 | **EQ-5D**, ZBPI | Y/N (not available) |  |
| Drolet et al. <sup>26</sup> | 2010 | Canada* | ≥50 | **EQ-5D**, SF-12 | **Y/Y** |  |
| Duracinsky et al. <sup>16</sup> | 2014 | France | ≥50 | SF-12, HADS | N |  |
| Gater et al. <sup>11</sup> | 2014 | UK | ≥50 | **EQ-5D**, SF-36, ZBPI | Y/N<br>(2 time points per<br>individual) |  |
| Lieu et al. <sup>13</sup> | 2008 | US | ≥60 | Time trade-off | N | Y |
| Lionis et al. <sup>23</sup> | 2011 | Greece | ≥50 | Likert, Short Anxiety Screening Test | N |  |
| Matthews et al. <sup>21</sup> | 2019 | Italy | ≥50 | **EQ-5D**, ZBPI | Y/N (not available) |  |
| Mizukami et al. <sup>27</sup> | 2018 | Japan | ≥60 | **EQ-5D**, SF-12, McGill, ZBPI | Y/N (not available) |  |
| Oster et al. <sup>14</sup> | 2005 | US | ≥75 | **EQ-5D** | Y/N<br>(1 time point per<br>individual) | Y |
| Pickering et al. <sup>17</sup> | 2016 | France | ≥50 | SF-12, HADS | N |  |
| Rampakakis et al. <sup>31</sup> | 2017 | Argentina, Brazil, Costa Rica,<br>Mexico, Canada*, South Korea*,<br>Taiwan*, Thailand | ≥50 | **EQ-5D**, EQ-VAS, ZBPI | **Y/Y**<br>(Brazil cohort data not<br>available) |  |
| Rampakakis et al. <sup>30</sup> | 2021 | Colombia | ≥50 | **EQ-5D**, ZBPI | Y/N (not available) | Y |
| Scott et al. <sup>10</sup> | 2006 | UK | Any | **EQ-5D** | **Y/Y** | Y |
| Song et al. <sup>28</sup> | 2014 | South Korea* | ≥50 | **EQ-5D** | **Y/Y** |  |
| Torcel-Pagnon et al. <sup>20</sup> | 2017 | Italy | ≥50 | SF-12 | N |  |
| Tsai et al. <sup>29</sup> | 2015 | Taiwan* | ≥50 | **EQ-5D** | **Y/Y** |  |
| van Wijck et al. <sup>24</sup> | 2017 | Netherlands | ≥50 | **EQ-5D**, SF-12 | **Y/Y** |  |
| Volpi et al. <sup>18</sup> | 2008 | Italy | Any | Short Italian Questionnaire | N | Y |

\* Three cohorts from Rampakakis et al. <sup>30</sup> are also included in other publications in this table. EQ-5D: EuroQol EQ-5D; EQ-VAS: EuroQol visual analogue scale; HADS: hospital anxiety and depression scale; SF-12: 12-item short-form survey; SF-36: 36-item short-form survey; ZBPI: zoster brief pain inventory; ZIQ: zoster impact questionnaire; McGill: McGill Pain Questionnaire Present Pain Intensity

## References

1. van Oorschot D, Vroling H, Bunge E, Diaz-Decaro J, Curran D, Yawn B. A systematic literature review of herpes zoster incidence worldwide. Hum Vaccin Immunother. 2021;17(6):1714–1732. doi:10.1080/21645515.2020.1847582

2. Kawai K, Gebremeskel BG, Acosta CJ. Systematic review of incidence and complications of herpes zoster: Towards a global perspective. BMJ Open. 2014;4(6):e004833. doi:10.1136/bmjopen-2014-004833

3. Harpaz R, Ortega-Sanchez IR, Seward JF. Prevention of herpes zoster: recommendations of the Advisory Committee on Immunization Practices (ACIP). MMWR Recommendations and reports : Morbidity and mortality weekly report Recommendations and reports / Centers for Disease Control. 2008;57(RR-5).

4. Rabin R, De Charro F. EQ-5D: A measure of health status from the EuroQol Group. Ann Med. 2001;33(5):337–343. doi:10.3109/07853890109002087

5. Rampakakis E, Stutz M, Kawai K, et al. Association between work time loss and quality of life in patients with Herpes Zoster: A pooled analysis of the MASTER studies. Health Qual Life Outcomes. 2017;15(1):1–14. doi:10.1186/s12955-017-0588-x

6. Tsai TF, Yao CA, Yu HS, et al. Herpes zoster-associated severity and duration of pain, health-related quality of life, and healthcare utilization in TWN: A prospective observational study. Int J Dermatol. 2015;54(5):529–536. doi:10.1111/ijd.12484

7. Drolet M, Brisson M, Schmader KE, et al. The impact of herpes zoster and postherpetic neuralgia on health-related quality of life: A prospective study. CMAJ Canadian Medical Association Journal. 2010;182(16):1731–1736. doi:10.1503/cmaj.091711

8. Song H, Lee J, Lee M, et al. Burden of illness, quality of life, and healthcare utilization among patients with herpes zoster in South Korea: A prospective clinical-epidemiological study. International Journal of Infectious Diseases. 2014;20(1):23–30. doi:10.1016/j.ijid.2013.11.018

9. R Studio Team. A language and environment for statistical computing. R Foundation for Statistical Computing. 2021;3:https://www.R-project.org. http://www.r-project.org

10. Scott FT, Johnson RW, Leedham-Green M, Davies E, Edmunds WJ, Breuer J. The burden of Herpes Zoster: A prospective population based study. Vaccine. 2006;24(9):1308–1314. doi:10.1016/j.vaccine.2005.09.026

11. Gater A, Abetz-Webb L, Carroll S, Mannan A, Serpell M, Johnson R. Burden of herpes zoster in the UK: Findings from the zoster quality of life (ZQOL) study. BMC Infect Dis. 2014;14(1):1–14. doi:10.1186/1471-2334-14-402

12. Coplan PM, Schmader K, Nikas A, et al. Development of a measure of the burden of pain due to herpes zoster and postherpetic neuralgia for prevention trials: Adaptation of the brief pain inventory. Journal of Pain. 2004;5(6):344–356. doi:10.1016/j.jpain.2004.06.001

13. Lieu TA, Ray GT, Ortega-Sanchez IR, Kleinman K, Rusinak D, Prosser LA. Willingness to pay for a QALY based on community member and patient preferences for temporary health states associated with Herpes Zoster. Pharmacoeconomics. 2009;27(12):1005–1016. doi:10.2165/11314000-000000000-00000

14. Oster G, Harding G, Dukes E, Edelsberg J, Cleary PD. Pain, medication use, and health-related quality of life in older persons with postherpetic neuralgia: Results from a population-based survey. Journal of Pain. 2005;6(6):356–363. doi:10.1016/j.jpain.2005.01.359

15. Bouhassira D, Chassany O, Gaillat J, et al. Patient perspective on herpes zoster and its complications: An observational prospective study in patients aged over 50 years in general practice. Pain. 2012;153(2):342–349. doi:10.1016/j.pain.2011.10.026

16. Duracinsky M, Paccalin M, Gavazzi G, et al. ARIZONA study: Is the risk of post-herpetic neuralgia and its burden increased in the most elderly patients? BMC Infect Dis. 2014;14(1). doi:10.1186/1471-2334-14-529

17. Pickering G, Gavazzi G, Gaillat J, Paccalin M, Bloch K, Bouhassira D. Is herpes zoster an additional complication in old age alongside comorbidity and multiple medications? Results of the post hoc analysis of the 12-month longitudinal prospective observational ARIZONA cohort study. BMJ Open. 2016;6(2):e009689. doi:10.1136/bmjopen-2015-009689

18. Volpi A, Gatti A, Pica F, Bellino S, Marsella LT, Sabato AF. Clinical and psychosocial correlates of post-herpetic neuralgia. J Med Virol. 2008;80(9):1646–1652. doi:10.1002/jmv.21254

19. Bricout H, Perinetti E, Marchettini P, et al. Burden of herpes zoster-associated chronic pain in Italian patients aged 50 years and over (2009-2010): A GP-based prospective cohort study. BMC Infect Dis. 2014;14(1). doi:10.1186/s12879-014-0637-6

20. Torcel-Pagnon L, Bricout HD, Bertrand I, et al. Impact of Underlying Conditions on Zoster-Related Pain and on Quality of Life Following Zoster. Journals of Gerontology - Series A Biological Sciences and Medical Sciences. 2017;72(8):1091–1097. doi:10.1093/gerona/glw189

21. Matthews S, De Maria A, Passamonti M, et al. The economic burden and impact on quality of life of herpes zoster and postherpetic neuralgia in individuals aged 50 years or older in Italy. Open Forum Infect Dis. 2019;6(2):ofz007. doi:10.1093/ofid/ofz007

22. Curran D, Schmidt-Ott R, Schutter U, Simon J, Anastassopoulou A, Matthews S. Impact of herpes zoster and postherpetic neuralgia on the quality of life of Germans aged 50 or above. BMC Infect Dis. 2018;18(1). doi:10.1186/s12879-018-3395-z

23. Lionis CD, Vardavas CI, Symvoulakis EK, et al. Measuring the burden of herpes zoster and post herpetic neuralgia within primary care in rural Crete, Greece. BMC Fam Pract. 2011;12. doi:10.1186/1471-2296-12-136

24. van Wijck AJM, Aerssens YR. Pain, Itch, Quality of Life, and Costs after Herpes Zoster. Pain Practice. 2017;17(6):738–746. doi:10.1111/papr.12518

25. Díez-Domingo J, Curran D, Cambronero M del R, Garcia-Martinez JA, Matthews S. Economic Burden and Impact on Quality of Life of Herpes Zoster in Spanish Adults Aged 50 Years or Older: A Prospective Cohort Study. Adv Ther. 2021;38(6):3325–3341. doi:10.1007/s12325-021-01717-7

26. Drolet M, Brisson M, Schmader KE, et al. The impact of herpes zoster and postherpetic neuralgia on health-related quality of life: A prospective study. CMAJ Canadian Medical Association Journal. 2010;182(16):1731–1736. doi:10.1503/cmaj.091711

27. Mizukami A, Sato K, Adachi K, et al. Impact of Herpes Zoster and Post-Herpetic Neuralgia on Health-Related Quality of Life in Japanese Adults Aged 60 Years or Older: Results from a Prospective, Observational Cohort Study. Clin Drug Investig. 2018;38(1):29–37. doi:10.1007/s40261-017-0581-5

28. Song H, Lee J, Lee M, et al. Burden of illness, quality of life, and healthcare utilization among patients with herpes zoster in South Korea: A prospective clinical-epidemiological study. International Journal of Infectious Diseases. 2014;20(1):23–30. doi:10.1016/j.ijid.2013.11.018

29. Tsai TF, Yao CA, Yu HS, et al. Herpes zoster-associated severity and duration of pain, health-related quality of life, and healthcare utilization in TWN: A prospective observational study. Int J Dermatol. 2015;54(5):529–536. doi:10.1111/ijd.12484

30. Rampakakis E, Stutz M, Monsanto-Planadeball HA, et al. Prospective observational study evaluating the disease burden of HZ in Colombia. Acta Medica Colombiana. 2021;46(3):11–18. http://www.scielo.org.co/scielo.php?script=sci_arttext&pid=S0120-24482021000300011&lng=en&nrm=iso&tlng=en%0Ahttp://www.scielo.org.co/scielo.php?script=sci_abstract&pid=S0120-24482021000300011&lng=en&nrm=iso&tlng=en

31. Rampakakis E, Stutz M, Kawai K, et al. Association between work time loss and quality of life in patients with Herpes Zoster: A pooled analysis of the MASTER studies. Health Qual Life Outcomes. 2017;15(1):1–14. doi:10.1186/s12955-017-0588-x

32. Oster G, Harding G, Dukes E, Edelsberg J, Cleary PD. Pain, medication use, and health-related quality of life in older persons with postherpetic neuralgia: Results from a population-based survey. Journal of Pain. 2005;6(6):356–363. doi:10.1016/j.jpain.2005.01.359

33. Rampakakis E, Pollock C, Vujacich C, et al. Economic Burden of Herpes Zoster (“culebrilla”) in Latin America. International Journal of Infectious Diseases. 2017;58:22–26. doi:10.1016/j.ijid.2017.02.021

34. National Institute for Health and Care Excellence (NICE). NICE health technology evaluation topic selection: the manual. 2022. Accessed July 25, 2024. www.nice.org.uk/process/pmg37

35. van Hoek AJ, Gay N, Melegaro A, Opstelten W, Edmunds WJ. Estimating the cost-effectiveness of vaccination against herpes zoster in England and Wales. Vaccine. 2009;27(9):1454–1467. doi:10.1016/j.vaccine.2008.12.024

36. Sollie M, Jepsen P, Sørensen JA. Patient-reported quality of life in patients suffering from acute herpes zoster—a systematic review with meta-analysis. Br J Pain. 2022;16(4):404–419. doi:10.1177/20494637211073050

37. Giannelos N, Ng C, Curran D. Cost-effectiveness of the recombinant zoster vaccine (RZV) against herpes zoster: An updated critical review. Hum Vaccin Immunother. 2023;19(1). doi:10.1080/21645515.2023.2168952

38. Sprangers MAG, Schwartz CE. Integrating response shift into health-related quality of life research: A theoretical model. Soc Sci Med. 1999;48(11):1507–1515. doi:10.1016/S0277-9536(99)00045-3

39. Oort FJ, Visser MRM, Sprangers MAG. Formal definitions of measurement bias and explanation bias clarify measurement and conceptual perspectives on response shift. J Clin Epidemiol. 2009;62(11):1126–1137. doi:10.1016/j.jclinepi.2009.03.013

40. Hillebrand K, Bricout H, Schulze-Rath R, Schink T, Garbe E. Incidence of herpes zoster and its complications in Germany, 2005-2009. Journal of Infection. 2015;70(2):178–186. doi:10.1016/j.jinf.2014.08.018

41. Schröder C, Enders D, Schink T, Riedel O. Incidence of herpes zoster amongst adults varies by severity of immunosuppression. J Infect. 2017;75(3):207–215. doi:10.1016/J.JINF.2017.06.010

42. Rommelaere M, Maréchal C, Yombi JC, Goffin E, Kanaan N. Disseminated varicella zoster virus infection in adult renal transplant recipients: Outcome and risk factors. Transplant Proc. 2012;44(9):2814–2817. doi:10.1016/j.transproceed.2012.09.090

43. Scott FT, Johnson RW, Leedham-Green M, Davies E, Edmunds WJ, Breuer J. The burden of Herpes Zoster: A prospective population based study. Vaccine. 2006;24(9):1308–1314. doi:10.1016/j.vaccine.2005.09.026

44. van Wijck AJM, Aerssens YR. Pain, Itch, Quality of Life, and Costs after Herpes Zoster. Pain Practice. 2017;17(6):738–746. doi:10.1111/papr.12518

45. Gater A, Abetz-Webb L, Carroll S, Mannan A, Serpell M, Johnson R. Burden of herpes zoster in the UK: Findings from the zoster quality of life (ZQOL) study. BMC Infect Dis. 2014;14(1):1–14. doi:10.1186/1471-2334-14-402

