## Supplementary material 2 for "Health-related quality of life with herpes zoster: systematic review and analysis of individual patient data from 9 cohorts"

### Supplementary information 2. supporting methods and results

### A Systematic review

#### A.1 Initial search

PubMed and Web of Science were systematically searched as follows:

**Pubmed**

- shingles OR zoster (title/abstract)
- burden OR qaly OR utility OR quality of life (title/abstract)
- 1.1.2009 to 31.10.2024 (publication date)
- English (language)

**Web of science**

- shingles OR zoster (topic)
- burden OR qaly OR utility OR "quality of life" (topic)
- 1.1.2009 to 31.10.2024 (publication date)
- English (language)

#### A.2 Inclusion/exclusion criteria

Abstracts for peer-reviewed studies concerning HZ were selected for full-text assessment if they conformed to any of the following criteria:

1. Reported a HRQoL measure;
2. Described cost-utility analyses (to find new utility estimates if they were used);
3. Described burden or epidemiology studies that included economic outcomes (e.g. costs), or presented a patient-reported outcome or a psychosocial outcome;
4. Reviewed the literature making explicit reference to cost-utility analyses, economic analysis, patient-reported outcomes, psychosocial outcomes or HRQoL.

Studies that lacked representative samples were excluded, such as:

1. Case report
2. Single centre study
3. Study conducted in specialised healthcare centres
4. Clinical trials

Studies were grouped by study type for full-text screening.

**Empirical studies** Apart from the criteria above, we included the observational studies that provided a novel (containing data not published in other studies) estimate of HRQoL with HZ, whether at one or multiple time points.

**Health-economic/epidemiological studies with modelling** We extracted the source studies for the parameters of the QALY related to HZ from the modelling studies. Without restricting the publication date, all sources were screened as empirical studies.

**Review studies** We iterated through studies from the reviews for empirical studies, health-economic studies, and burden estimates. The records that met our criteria were included if not included before.

### B Data description and data pre-processing

#### B.1 Calculation of time since rash onset

To understand the changes of EQ-5D since the rash onset. We used the number of days as a key covariate if available. For those with time points not exactly recorded, the scheduled time for the completion of the EQ-5D questionnaires was applied as an alternative. For Scott et al.[14], the number of days since the rash onset was calculated by subtracting the rash onset date from the visit date. For the patient cohorts in Rampakakis et al. [1], only the number of days since rash onset was recorded at the first visit. The number of days since rash onset at subsequent visits for these patient cohorts was calculated by adding the time-point at which the questionnaire was due to be completed (e.g. 3 months after the first visit) to the number of days since rash onset recorded during the first visit. van Wijck et al. [2] recruited patients no more than 7 days after rash onset. At the first visit, 4 days were assumed to have passed since the rash onset. The number of days since rash onset at subsequent visits for van Wijck et al. [2] was calculated by adding the time point at which the questionnaire was due to be completed (e.g. 3 months after the first visit) to the 4 days since rash onset assumed at the first visit. Drolet et al. [3] had estimated the days since rash onset until each EQ-5D measurement, and we were able to use the estimator directly.

#### B.2 Data description

In total, there were 9,810 data points of 1,345 subjects in 9 studies fetched. The measures per subject ranged between 4 and 10 times. There are 66 subjects aged below 50 years old, 467 aged 50-59, 425 aged 60-69, 274 aged 70-79, and 113 aged 80+. The EQ-5D score ranged between -0.594 and 1. There were 5,066 (51.6%) perfect health (EQ-5D = 1) data points; 341 points with perfect health had non-perfect health following.

### C Exploratory data analysis

#### C.1 Overview

The dynamics of EQ-5D scores provided two types of information: the duration with HZ-related disutility and the values of EQ-5D scores. We intended to model them separately. We applied a stratified sampling on the full data for sampling 25% of individual patients for each cohort. The final model will be built with Bayesian approaches. However, considering the computational time, we might start with Frequentist's approaches at the earlier stage. For the Bayesian model selection, we employed the Watanabe–Akaike information criterion (WAIC) and Leave-one-out cross-validation (LOO) for model selection; for Frequentist's approaches, we employed the Akaike information criterion (AIC) and mean squared error (MSE).

#### C.2 Disutility period, time-to recovery from HZ

The disutility period is the time-to-event data, so we applied survival analysis. Without knowing the baseline QALY, we defined the QALY loss as any EQ-5D value that is below 1. There are two types of data endpoints. If there is a clear endpoint with no QALY loss in the last measurements, the endpoints will be within the intervals of the last point with QALY loss and the time of the next measurement. If the QALY loss remains at the end of the data series, the endpoints will be after the last observations, which are the ordinary right censoring. The explorative analyses used R with *survival*, *survstan*, and *rstan* packages.

**Step 1-1** We explored the functions of age. For convenience, we took the mid-points of the intervals as the time-to-event proxies for those non-censored. We employed Cox's proportional hazard model which does not need the baseline hazard function being fully parameterised.

**Step 1-2** We iterated through *Weibull*, *Gamma*, *log-normal*, and *Gompertz* distributions for the baseline hazard. The AICs and MSEs are as follows.

*Log-normal* and *Gompertz* failed to converge well during model-fitting since the hyper-parameters trend to overflow numerically. The *Weibull* and *gamma* fits performed better than the others while they performed similarly.

**Step 2** In this step, we ran fully structured models on the training data. As both *Weibull* and *gamma* distributions were selected, we added the *exponential* distribution as the special cases for them as well. See the supplementary material of methods for the fully detailed model structure.

By assessing the WAICs and LOOs, the data exponential distribution performed better than the other two. From the posterior distributions of the hyperparameters, both the *Weibull* and *gamma* distributions were reduced to *exponential* distribution. As a result, we finally utilised an exponential model with a hazard ratio linking to age as the model for the disutility period.

#### C.3 EQ-5D scores, QALY with HZ

Firstly, we identified three clusters in the EQ-5D scores before the end of disutility periods as Fig-S4 showed. Since there are clear boundaries between the clusters, we used the K-means to classify the data points by temporally well, mild discomfort, and severe discomfort. Within each cluster, we identified models separately and the proportions of the clusters are modelled separately as well. In total, there are four sub-models for the dynamics of EQ-5D scores: (1) the dynamics of mild discomfort, (2) the dynamics of severe discomfort, (3) the probability of temporal well-off, and (4) the probability of severe discomfort within those has QALY-loss.

In general, the patient's age and time since rash onset are the key predictors to be tested. As it is a dataset with repeat measurements, we considered the random effects for patients and source studies. For computation time concerns, we started with the Frequentist's approach like the time-to-recovery data via *lme4* package in R. The models that failed to converge during modelling or have identifiability issues will be removed before model comparison. The model function expression fellows the *lme4* formula in R. Based on the model specified by the Frequentist's approach, we transformed to Bayesian approaches via stan. We explored reduced models, which step-wisely removed age- or time-related predictors

The models were built from the basic models with temporal patterns and random effects. Then, we tried to relate the model to patients' age, squared age, or higher order if needed. There are always intercept-only models as null models. The candidate model components were as follows.

**Basic models (intercepts):**

- Y ∼ 1
- Y ∼ 1 + (1 | patient)
- Y ∼ 1 + (1 | cohort)
- Y ∼ 1 + (1 | patient) + (1 | cohort)

**Temporal patterns:**

- no time-specific term
- + time
- + (time | patient)
- + (time | cohort)
- + (time | patient) + (time | cohort)
- + d15
- + d15 + d30
- + d15 + d30 + d45
- + d15 + d30 + d45 + d60

**Age patterns:**

- no age-specific term
- + age
- + age + age $^2$

As the disutility period, the exploratory analyses were executed on 25% subset patients. The details of the analyses are trivial, so we only listed the key findings here. The detailed process can be found in the project GitHub repository of this study.

##### C.3.1 Sub-model 1: mild discomfort

- For time-dependency, random slopes for patients and studies performed better than time as a covariate with random effects for patients and studies.
- Adding an age pattern as a predictor reduced the performance
- Random effects by cohort-specific slopes of time explained most of the variation
- Best fit: y $\sim$ 1 + (time | cohort)
- Prediction with fixed-effects: y $\sim$ 1

##### C.3.2 Sub-model 2: severe discomfort

- Similar model specification as the mild discomfort
- Best fit: y $\sim$ 1 + (time | cohort)
- Prediction with fixed-effects: y $\sim$ 1

##### C.3.3 Sub-model 3: probability of temporally well (logistic regression)

- Linear time-trends were unfavourable and had low performance as the residuals are skewed after months since rash onset.
- The patient-specific random effect caused identifiability issues.
- Discrete 15- and 30-day dummy variables provide better goodness of fit than the other temporal patterns.
- Linear age pattern preferred. However, the respective coefficients are not significantly greater/smaller than zero.
- The Bayesian approach suggested removing age-related terms.
- Best fit: logit(y) $\sim$ d15 + d30 + (1 | cohort)
- Prediction with fixed-effects: logit(y) $\sim$ d15 + d30

##### C.3.4 Sub-model 4: the probability of mild discomfort among those has QALY loss (logistic regression)

- Linear time-trend has low performance as the residuals are biased months after rash onset.
- Discrete 15- and 30-day dummy variables provide better goodness of fit than the other temporal patterns.
- Quadratic polynomial function of age preferred.
- The Bayesian approach suggested removing age-related terms.
- Best fit: logit(y) $\sim$ d15 + d30 + (1 | cohort)
- Prediction with fixed-effects: logit(y) $\sim$ d15 + d30

### D Empirical models

#### D.1 Empirical model of disutility period of HZ

##### D.1.1 Data

There are two groups of patients in the data. The first group ($g_1$) has events between two observation points, ie, $t_i$, and $z_i$, where $i$ indicates the $i$th patient. The other ($g_2$) has the last observation time, $t_i$, with the event time censored.

##### D.1.2 Statistical model

Given a cumulative density function of the time-to-event distribution, $F(t)$, the likelihood function of the model is composed according to the exploratory analyses in Appendix C. For each patient $i$ in the first group with $t_i$ and $z_i$, the probability that the event occurs between $t_i$ and $z_i$ is

$$F_\alpha(z_i) - F_\alpha(t_i)$$

where

$$F_\alpha(t) = 1 - exp(-r_0 e^{\alpha\beta_\alpha} t)$$

for specified exponential distribution

In the second group, the data inform the likelihood function by the probability that the event happens after $t_i$. That is, the complementary cumulative density

$$1 - F_\alpha(t_i)$$

The log-likelihood function is written as:

$$log(l) = \sum_{i \in g1} log(F_\alpha(z_i) - F_\alpha(t_i)) + \sum_{i \in g2} log(1 - F_\alpha(t_i))$$

#### D.2 Empirical model of HRQoL with HZ

##### D.2.1 Data

The data are repeatly measured quality of life ($q_{ij}$) for the $j$th measure of the patient $i$. The number of patients $N$, the number of measures for each patient $i$ is used, and each patient $i$ has a source cohort $cohort_i$

##### D.2.2 Statistical model

Following a mixture model with the sub-models specified in the exploratory analyses (Appendix C), the full model is written as:

$$
\begin{aligned}
q_{ij} &\sim mixture(\pi^z_{ij}, 1, \pi^m_{ij}, N(\mu^m_{ij}, \epsilon^m)I(-\infty, 1), \pi^s_{ij}, N(\mu^s_{ij}, \epsilon^s)I(-\infty, 1) \\
\mu^m_{ij} &= \beta^1_0 + t_{ij}\beta^1_{cohort} \\
\mu^s_{ij} &= \beta^2_0 + t_{ij}\beta^2_{cohort} \\
logit(\pi^z_{ij}) &= \beta^3_0 + I[t_{ij} > 15]\beta^3_{d15} + I[t_{ij} > 30]\beta^3_{d30} + \beta^3_{cohort} \\
logit(\frac{\pi^m_{ij}}{\pi^m_{ij} + \pi^s_{ij}}) &= \beta^4_0 + I[t_{ij} > 15]\beta^4_{d15} + I[t_{ij} > 30]\beta^4_{d30} + \beta^4_{cohort} \\
\pi^z_{ij} + \pi^m_{ij} + \pi^s_{ij} &= 1
\end{aligned}
$$

**Notations:**

- $mixture(p_1, d_1, ..., p_n, d_n)$: mixture model combining distributions from $d_1$ to $d_n$ via proportions from $p_1$ to $p_n$
- $N(\mu, \sigma)I(-\infty, 1)$: normal distribution with mean ($\mu$), standard deviation ($\sigma$), capped by 1
- $\pi^z_{ij}$: proportion of cluster $\in \{z, m, s\}$**temporally well, mild discomfort, severe discomfort**
- $g_k(.)$: linear function for submodel $k$ as specified in Appendix C

#### D.3 Posterior distribution

Table 1: Posterior distribution.

| Parameter | mean | sd | 2.5% | 25% | 50% | 75% | 97.5% | ESS | Rhat |
|---|---|---|---|---|---|---|---|---|---|
| Disutility period of HZ | | | | | | | | | |
| $r_0$ | 5.40 | 0.92 | 3.71 | 4.76 | 5.36 | 5.97 | 7.39 | 381.47 | 1.01 |
| $\beta_\alpha$ | -0.01 | 0.00 | -0.02 | -0.01 | -0.01 | -0.01 | -0.01 | 379.44 | 1.01 |
| HRQoL with HZ, sub-model 1: mild discomfort | | | | | | | | | |
| $\beta_0^1$ | 0.74 | 0.00 | 0.73 | 0.74 | 0.74 | 0.74 | 0.74 | 1983.48 | 1.00 |
| HRQoL with HZ, sub-model 2: severe discomfort | | | | | | | | | |
| $\beta_0^2$ | 0.10 | 0.01 | 0.08 | 0.09 | 0.10 | 0.10 | 0.11 | 3155.82 | 1.00 |
| HRQoL with HZ, sub-model 3: Pr(temporally well) | | | | | | | | | |
| $\beta_0^3$ | -1.28 | 0.16 | -1.59 | -1.38 | -1.28 | -1.18 | -0.97 | 3534.11 | 1.00 |
| $\beta_{d15}^3$ | -0.80 | 0.44 | -1.66 | -1.10 | -0.79 | -0.49 | 0.03 | 2853.90 | 1.00 |
| $\beta_{d30}^3$ | -1.03 | 0.32 | -1.67 | -1.23 | -1.02 | -0.81 | -0.43 | 2470.35 | 1.00 |
| HRQoL with HZ, sub-model 4: Pr(mild discomfort\|not temporally well) | | | | | | | | | |
| $\beta_0^4$ | 2.36 | 0.09 | 2.18 | 2.30 | 2.36 | 2.42 | 2.53 | 1839.06 | 1.00 |
| $\beta_{d15}^4$ | -0.76 | 0.10 | -0.96 | -0.83 | -0.75 | -0.69 | -0.56 | 2830.78 | 1.00 |
| $\beta_{d30}^4$ | -0.56 | 0.12 | -0.79 | -0.64 | -0.56 | -0.48 | -0.33 | 2183.06 | 1.00 |

ESS: effective sample size

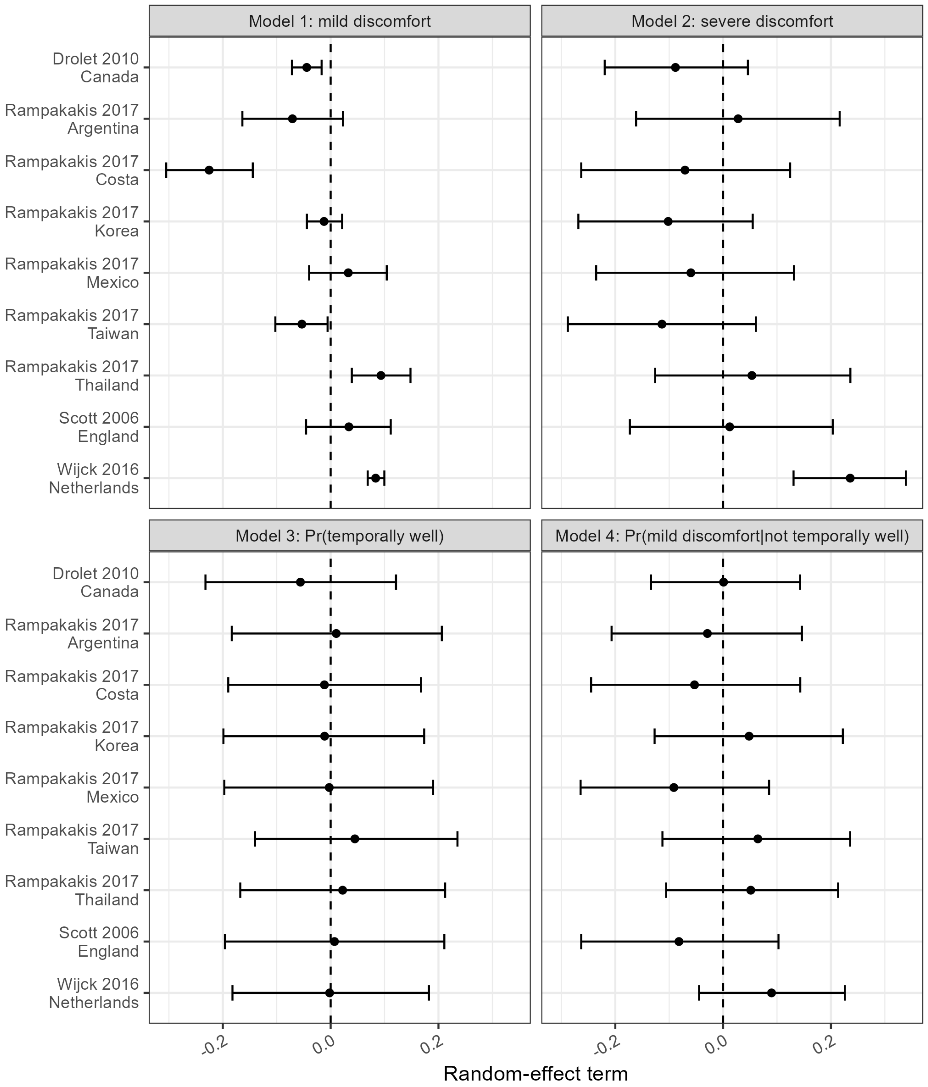

Figure 1: Posterior distribution of random-effect terms by model. Cohort-specific slopes for Model 1 and Model 2; cohort-specific intercepts for Model 3 and Model 4

### E Calculation of QALY loss due to HZ

The QALY loss due to HZ counts the difference between the HRQoL with HZ and the baseline health status across the disutility period. For the decision-making purpose, a discounting factor can be applied to convert the QALY loss to the present value considering the time preference.

#### E.1 Definitions and notations

**Time-related**

$t$**:** time since the onset of HZ rash.

$\alpha$**:** age of the onset of HZ rash.

$x$**:** age ($= \alpha + t$)

**HRQoL parameters**

$q^m$**:** quality of life, mild discomfort.

$q^s$**:** quality of life, severe discomfort.

$p^m_\alpha$**:** probability of being mildly affected by HZ.

$p^s_\alpha$**:** probability of being severely affected by HZ.

$p^z_\alpha$**:** probability of being temporally well.

$r_0$**:** baseline rate of recovery.

$\beta_\alpha$**:** log rate ratio of recovery rate for age (ref = 0).

**External inputs**

$H_x$**:** baseline HRQoL at age $x$, i.e. population norm.

$\delta$**:** discounting factor, time preference.

#### E.2 Calculations

**Disutility period** According to our statistical analysis, the duration of the disutility period is captured by an exponential distribution. The rate of HZ recovery (from the disutility period) is associated with age ($\alpha$):

$$log(r_\alpha) = log(r_0) + \alpha\beta_\alpha$$

For an exponential distribution, the survival function, which is the probability of staying at the disutility period, is:

$$S_\alpha(t) = e^{-r_\alpha t}$$

**HRQoL with HZ** The HRQoL with HZ is captured by a mixture distribution with (1) HRQoL mildly affected by HZ, (2) HRQoL severely affected by HZ, and (3) temporally well. With the model, the expected HRQoL is composed of:

$$q_\alpha(t) = p^m_\alpha q^m + p^s_\alpha q^s + p^z_\alpha$$

Regarding $H_{\alpha+t}$, baseline health status, as the upper bound of HRQoL with HZ, the expected HRQoL therefore becomes:

$$q_\alpha(t) = p^m_\alpha min(q^m, H_{\alpha+t}) + p^s_\alpha min(q^s, H_{\alpha+t}) + p^z_\alpha H_{\alpha+t}$$

**QALY loss due to HZ** The HRQoL after the onset of HZ rash is the HRQoL with and without HZ by their respective probabilities, $S_\alpha(t)$ and $1 - S_\alpha(t)$. That is,

$$q_\alpha(t)S_\alpha(t) + H_{\alpha+t}\big(1 - S_\alpha(t)\big)$$

Then, the loss in HRQoL is the difference between HRQoL with HZ and HRQoL in the baseline health status:

$$H_{\alpha+t} - \Big(q_\alpha(t)S_\alpha(t) + H_{\alpha+t}\big(1 - S_\alpha(t)\big)\Big) = \Big(H_{\alpha+t} - q_\alpha(t)\Big)S_\alpha(t)$$

The overall QALY loss cumulates the HRQoL from the rash onset to the end of the disutility period. Also, during the period, a time preference $\delta$ can be applied. Therefore, the present value of QALY loss due to HZ, $L_\alpha$, is

$$L_\alpha = \int_{t=0}^{\infty} \Big(H_{\alpha+t} - q_\alpha(t)\Big)S_\alpha(t)e^{-\delta t}dt$$

A special case uses perfect health as the baseline health status, i.e., $H_{a+t} = 1$. The $q_\alpha(.)$ is not time-varying as a result. The QALY loss can be simplified as

$$\begin{aligned} L_\alpha &= \int_{t=0}^{\infty} \big(1 - q_\alpha(.)\big)S_\alpha(t)e^{-\delta t}dt \\ &= \big(1 - q_\alpha(.)\big)\int_{t=0}^{\infty} e^{-r_\alpha t}e^{-\delta t}dt \\ &= \big(1 - q_\alpha(.)\big) \times \frac{1}{r_\alpha + \delta} \end{aligned}$$

### F Sources for population norms

We fetched the values of population norms from EuroQol suggested resources (https://euroqol.org/information-and-support/resources/population-norms/). We mapped included studies and the population norms sources with the following considerations:

1. Geographical relevance of the study site
2. Gross domestic product per capita.
3. Range of age
4. Date of publication

| Study | Matched setting | Source |
|---|---|---|
| Canada[1, 3] | Canada | Poder et al.[4] |
| Argentina[1] | Argentina | Janssen et al.[5] |
| Costa Rica[1] | Brazil | Santos et al.[6] |
| Korea [1, 7] | Korea | Janssen et al.[5] |
| Mexico[1] | Brazil | Santos et al.[6] |
| Taiwan [1, 8] | Korea | Janssen et al.[5] |
| Thailand [1] | Thailand | Janssen et al.[5] |
| UK [9] | UK | McNamara et al.[10] |
| Netherlands [2] | Netherlands | Janssen et al.[5] |

Table 2: Sources of population norms

### G Alternative results with baseline EQ-5D

#### G.1 Estimates based on the pre-HZ EQ-5D scores

There were 54% of samples (722/1,335) that had EQ-5D scores surveyed retrospectively after HZ rash onset. We presented the alternative results using the baseline EQ-5D scores as the upper limit for each patients. The end points of disutility periods were defined accordingly and EQ-5D scores were bounded as well.

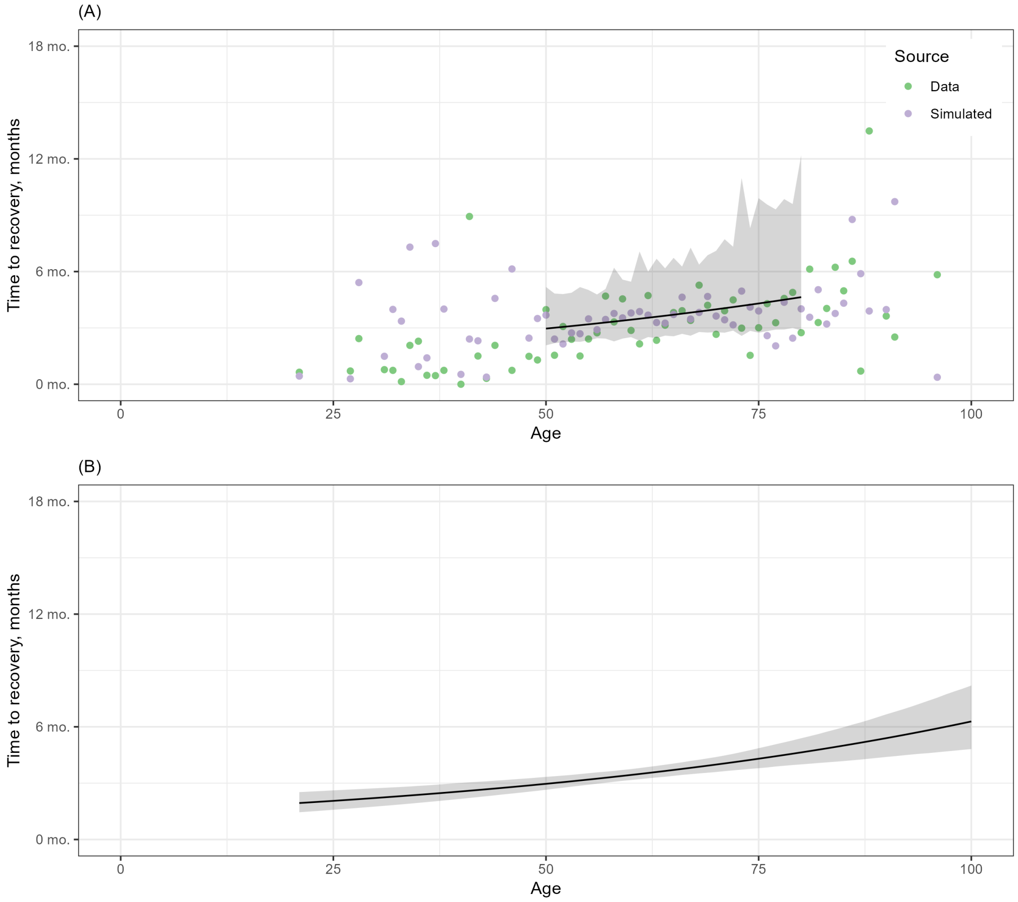

Figure 2: **Disutility period with pre-HZ baseline.** (A) Average disutility period by age. Red points for mean of the imputed recovery time, blue points for mean of the simulated recovery time given the same population size as the data. (B) 95% credible intervals of disutility periods by age.

#### G.2 Results based on population norms

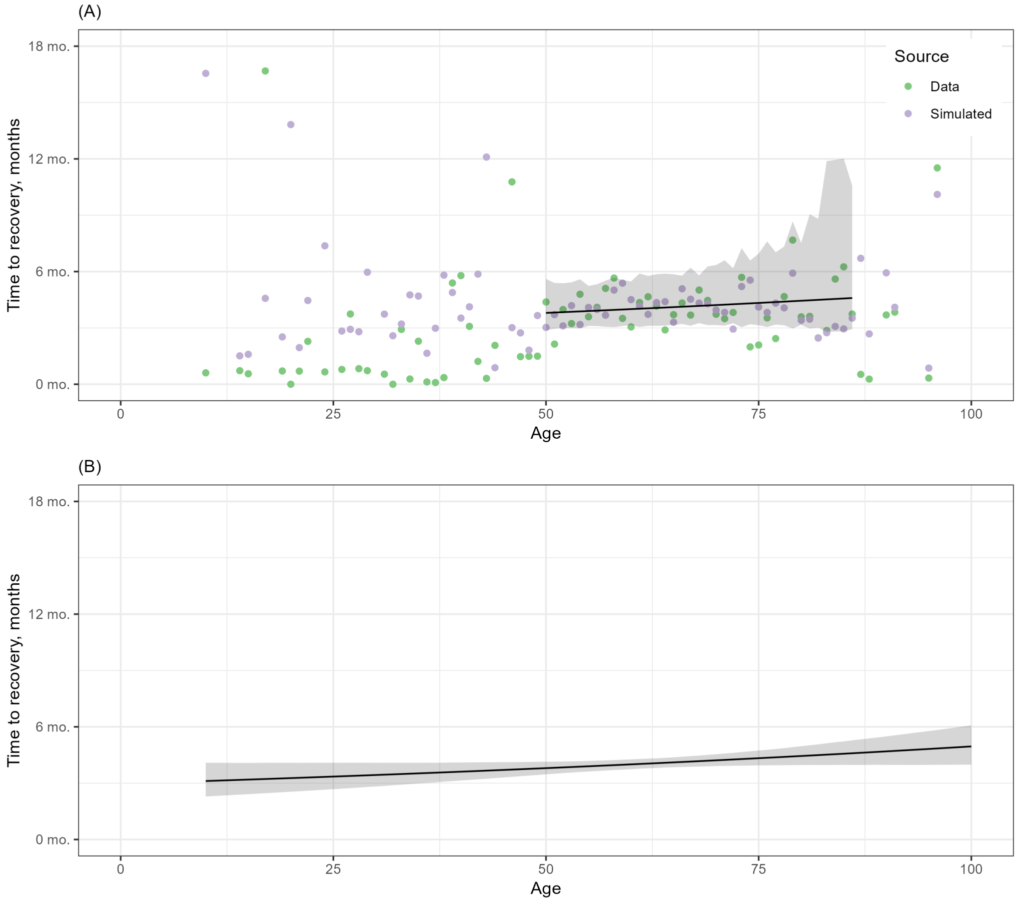

Figure 3: **Disutility period with population norms as endpoints.** (A) Average disutility period by age. Red points for mean of the imputed recovery time, blue points for mean of the simulated recovery time given the same population size as the data. (B) 95% credible intervals of disutility periods by age.

| study | Age | QALY loss<br>no discounting | QALY loss<br>1.5% discounting | QALY loss<br>3.5% |
|---|---|---|---|---|
| Drolet et al. 2010<br>Canada | 50 | 0.040 (0.035 - 0.044) | 0.039 (0.035 - 0.044) | 0.039 (0.035 - 0.043) |
| | 60 | 0.043 (0.039 - 0.047) | 0.043 (0.039 - 0.047) | 0.042 (0.039 - 0.047) |
| | 70 | 0.047 (0.042 - 0.051) | 0.046 (0.042 - 0.051) | 0.046 (0.042 - 0.051) |
| | 80 | 0.047 (0.041 - 0.053) | 0.046 (0.041 - 0.052) | 0.046 (0.041 - 0.052) |
| | 90 | 0.046 (0.039 - 0.054) | 0.046 (0.039 - 0.054) | 0.046 (0.039 - 0.054) |
| Rampakakis et al. 2017<br>Argentina | 50 | 0.050 (0.046 - 0.056) | 0.050 (0.045 - 0.055) | 0.050 (0.045 - 0.055) |
| | 60 | 0.046 (0.042 - 0.051) | 0.046 (0.042 - 0.050) | 0.046 (0.042 - 0.050) |
| | 70 | 0.036 (0.032 - 0.040) | 0.035 (0.032 - 0.039) | 0.035 (0.032 - 0.039) |
| | 80 | 0.025 (0.022 - 0.029) | 0.025 (0.022 - 0.029) | 0.025 (0.022 - 0.029) |
| | 90 | 0.024 (0.020 - 0.028) | 0.024 (0.020 - 0.028) | 0.023 (0.020 - 0.028) |
| Rampakakis et al. 2017<br>Costa Rica | 50 | 0.033 (0.030 - 0.037) | 0.033 (0.030 - 0.037) | 0.033 (0.029 - 0.037) |
| | 60 | 0.028 (0.025 - 0.032) | 0.028 (0.025 - 0.031) | 0.028 (0.025 - 0.031) |
| | 70 | 0.029 (0.025 - 0.032) | 0.029 (0.025 - 0.032) | 0.028 (0.025 - 0.032) |
| | 80 | 0.030 (0.026 - 0.034) | 0.030 (0.026 - 0.034) | 0.030 (0.026 - 0.034) |
| | 90 | 0.032 (0.027 - 0.038) | 0.031 (0.026 - 0.037) | 0.031 (0.026 - 0.037) |
| Rampakakis et al. 2017<br>Korea | 50 | 0.064 (0.058 - 0.071) | 0.064 (0.058 - 0.070) | 0.064 (0.058 - 0.070) |
| | 60 | 0.049 (0.045 - 0.054) | 0.049 (0.045 - 0.053) | 0.049 (0.045 - 0.053) |
| | 70 | 0.038 (0.034 - 0.042) | 0.038 (0.034 - 0.042) | 0.038 (0.034 - 0.042) |
| | 80 | 0.036 (0.032 - 0.041) | 0.036 (0.032 - 0.041) | 0.036 (0.031 - 0.041) |
| | 90 | 0.038 (0.032 - 0.045) | 0.038 (0.032 - 0.045) | 0.038 (0.032 - 0.044) |
| Rampakakis et al. 2017<br>Mexico | 50 | 0.033 (0.030 - 0.037) | 0.033 (0.030 - 0.037) | 0.033 (0.029 - 0.037) |
| | 60 | 0.028 (0.025 - 0.032) | 0.028 (0.025 - 0.031) | 0.028 (0.025 - 0.031) |
| | 70 | 0.029 (0.025 - 0.032) | 0.029 (0.025 - 0.032) | 0.028 (0.025 - 0.032) |
| | 80 | 0.030 (0.026 - 0.034) | 0.030 (0.026 - 0.034) | 0.030 (0.026 - 0.034) |
| | 90 | 0.032 (0.027 - 0.038) | 0.031 (0.026 - 0.037) | 0.031 (0.026 - 0.037) |
| Rampakakis et al. 2017<br>Taiwan | 50 | 0.064 (0.058 - 0.071) | 0.064 (0.058 - 0.070) | 0.064 (0.058 - 0.070) |
| | 60 | 0.049 (0.045 - 0.054) | 0.049 (0.045 - 0.053) | 0.049 (0.045 - 0.053) |
| | 70 | 0.038 (0.034 - 0.042) | 0.038 (0.034 - 0.042) | 0.038 (0.034 - 0.042) |
| | 80 | 0.036 (0.032 - 0.041) | 0.036 (0.032 - 0.041) | 0.036 (0.031 - 0.041) |
| | 90 | 0.038 (0.032 - 0.045) | 0.038 (0.032 - 0.045) | 0.038 (0.032 - 0.044) |
| Rampakakis et al. 2017<br>Thailand | 50 | 0.020 (0.018 - 0.023) | 0.020 (0.018 - 0.023) | 0.020 (0.018 - 0.023) |
| | 60 | 0.020 (0.018 - 0.023) | 0.020 (0.018 - 0.023) | 0.020 (0.018 - 0.023) |
| | 70 | 0.020 (0.018 - 0.023) | 0.020 (0.018 - 0.023) | 0.020 (0.018 - 0.023) |
| | 80 | 0.020 (0.018 - 0.024) | 0.020 (0.018 - 0.024) | 0.020 (0.017 - 0.024) |
| | 90 | 0.021 (0.018 - 0.025) | 0.021 (0.018 - 0.025) | 0.021 (0.018 - 0.025) |
| Scott et al. 2006<br>United Kingdom | 50 | 0.043 (0.039 - 0.048) | 0.043 (0.039 - 0.048) | 0.043 (0.039 - 0.047) |
| | 60 | 0.040 (0.036 - 0.044) | 0.039 (0.036 - 0.043) | 0.039 (0.036 - 0.043) |
| | 70 | 0.033 (0.030 - 0.037) | 0.033 (0.029 - 0.037) | 0.033 (0.029 - 0.037) |
| | 80 | 0.023 (0.020 - 0.027) | 0.023 (0.020 - 0.027) | 0.023 (0.020 - 0.027) |
| | 90 | 0.023 (0.019 - 0.027) | 0.022 (0.019 - 0.027) | 0.022 (0.019 - 0.027) |
| van Wijck et al. 2016<br>The Netherlands | 50 | 0.060 (0.054 - 0.066) | 0.060 (0.054 - 0.066) | 0.060 (0.054 - 0.065) |
| | 60 | 0.058 (0.054 - 0.063) | 0.058 (0.053 - 0.063) | 0.058 (0.053 - 0.063) |
| | 70 | 0.054 (0.049 - 0.059) | 0.054 (0.049 - 0.059) | 0.054 (0.049 - 0.059) |
| | 80 | 0.048 (0.042 - 0.054) | 0.048 (0.042 - 0.054) | 0.048 (0.042 - 0.054) |
| | 90 | 0.044 (0.037 - 0.052) | 0.044 (0.037 - 0.052) | 0.044 (0.037 - 0.051) |

Table 3: QALY loss due to HZ by country-specific population norms.

### References

[1] Rampakakis E, Stutz M, Kawai K, Tsai TF, Cheong HJ, Dhitavat J, et al. Association between work time loss and quality of life in patients with Herpes Zoster: A pooled analysis of the MASTER studies. Health and Quality of Life Outcomes. 2017 jan;15(1):1-14. Available from: https://link.springer.com/article/10.1186/s12955-017-0588-x.

[2] van Wijck AJM, Aerssens YR. Pain, Itch, Quality of Life, and Costs after Herpes Zoster. Pain Practice. 2017 jul;17(6):738-46. Available from: https://onlinelibrary.wiley.com/doi/full/10.1111/papr.12518https://onlinelibrary.wiley.com/doi/abs/10.1111/papr.12518https://onlinelibrary.wiley.com/doi/10.1111/papr.12518.

[3] Drolet M, Brisson M, Schmader KE, Levin MJ, Johnson R, Oxman MN, et al. The impact of herpes zoster and postherpetic neuralgia on health-related quality of life: A prospective study. CMAJ Canadian Medical Association Journal. 2010 nov;182(16):1731-6. Available from: https://www.cmaj.ca/content/182/16/1731https://www.cmaj.ca/content/182/16/1731.abstract.

[4] Poder TG, Carrier N, Kouakou CRC. Quebec Health-Related Quality-of-Life Population Norms Using the EQ-5D-5L: Decomposition by Sociodemographic Data and Health Problems. Value in Health. 2020 2;23:251-9. Available from: https://www.sciencedirect.com/science/article/pii/S1098301519323526.

[5] Janssen MF, Szende A, Cabases J, Ramos-Goñi JM, Vilagut G, König HH. Population norms for the EQ-5D-3L: a cross-country analysis of population surveys for 20 countries. European Journal of Health Economics. 2019 3;20:205-16. Available from: https://link.springer.com/article/10.1007/s10198-018-0955-5.

[6] Santos M, Monteiro AL, Santos B. EQ-5D Brazilian population norms. Health and Quality of Life Outcomes. 2021 12;19:1-7. Available from: https://link.springer.com/articles/10.1186/s12955-021-01671-6https://link.springer.com/article/10.1186/s12955-021-01671-6.

[7] Song H, Lee J, Lee M, Choi WS, Choi JH, Lee MS, et al. Burden of illness, quality of life, and healthcare utilization among patients with herpes zoster in South Korea: A prospective clinical-epidemiological study. International Journal of Infectious Diseases. 2014 mar;20(1):23-30.

[8] Tsai TF, Yao CA, Yu HS, Lan CC, Chao SC, Yang JH, et al. Herpes zoster-associated severity and duration of pain, health-related quality of life, and healthcare utilization in TWN: A prospective observational study. International Journal of Dermatology. 2015 may;54(5):529-36. Available from: https://onlinelibrary.wiley.com/doi/full/10.1111/ijd.12484https://onlinelibrary.wiley.com/doi/abs/10.1111/ijd.12484https://onlinelibrary.wiley.com/doi/10.1111/ijd.12484.

[9] Scott FT, Johnson RW, Leedham-Green M, Davies E, Edmunds WJ, Breuer J. The burden of Herpes Zoster: A prospective population based study. Vaccine. 2006;24(9):1308-14. Available from: https://www.sciencedirect.com/science/article/pii/S0264410X05009801?casa_token=8Z6p0ZMESPkAAAAA:77tLZA_Jt9ZOLX24_p1C1PIANhKDAqPwuvj2ina4OwW-OOlTWdqhC-YdjlLVwbRmygO6G2Vc7HA.

[10] McNamara S, Schneider PP, Love-Koh J, Doran T, Gutacker N. Quality-Adjusted Life Expectancy Norms for the English Population. Value in Health. 2023 2;26:163-9. Available from: https://www.sciencedirect.com/science/article/pii/S1098301522021015.
